# Development and validation of a multiple-choice question-based delirium care knowledge quiz for critical care nurses

**DOI:** 10.1101/2021.01.16.21249923

**Authors:** Mu-Hsing Ho

## Abstract

**Aims:** To develop and psychometrically test a multiple choice questions (MCQs)-based quiz of delirium care knowledge for critical care nurses.

**Design:** Instrument development and psychometric evaluation study.

**Methods:** The development and validation process including two phases. Phase I focused on the quiz development, conducted by the following steps: (1) generated initial 20-item pool; (2) examined content validity and (3) face validity; (4) conducted pilot testing, data were collected from 217 critical care nurses via online survey during 01 October to 07 November, 2020; (5) performed item analysis and eliminated items based on the item difficulty and discrimination indices. The MCQs quiz was finalised through the development process. Then, phases II emphasised the quiz validation, to estimate the internal consistency, split-half and test-retest reliability, and construct validity using parallel analysis with the exploratory factor analysis (EFA).

**Results:** A final 16-item MCQs quiz was emerged from the item analysis. The Kuder– Richardson Formula 20 coefficient for the overall quiz showed good internal consistency (0.85), and the intraclass correlation coefficient with a 30-day interval also indicated that the questionnaire had satisfactory stability (0.96). The EFA confirmed appropriate construct validity for the quiz, four factors could explain the total variance of 60.87%.

**Conclusion:** This study developed the first MCQs quiz for delirium care knowledge and it is a reliable and valid tool that can be implemented to assess the level of delirium care knowledge.

**Impact:** This study offers an evidence-based quiz designed for future research and education purposes in delirium care that has significant implications for knowledge test by using MCQs in clinical practice.

## 1. INTRODUCTION

Delirium is the most common complication occurs among hospitalised patients, particularly in the intensive care unit (ICU). It is noted that delirium as a feature of COVID-19 may be increasingly found in critically ill patients who had severe infection and mechanically ventilated in ICU (British Geriatrics Society, European Delirium Association, & Royal College of Psychiatrists, 2020). This updated guidance on delirium care also highlighted the importance of delirium prevention, early detection, assessment and management. According to the clinical practice guidelines for the prevention and management of Pain, Agitation/Sedation, Delirium, Immobility, and Sleep Disruption (PADIS) in adult patients in the ICU, evidence of delirium risk factors, prediction, and assessment were summarised for healthcare provider (Devlin et al., 2018). To understand the level of updated delirium care knowledge among critical care nurses is crucial as they are the frontline care providers in ICU. To date, several instruments investigating the delirium knowledge among ICU healthcare professionals were developed. However, only few instruments targeting critical care nurses as main population were comprehensively validated and evaluated (Elliott, 2014; Monfared, Soodmand, & Ghasemzadeh, 2017; Öztürk Birge, Tel Aydın, & Salman, 2020). Most developed instruments utilised options in true/false design to assess the knowledge of delirium. No existing instrument adopted the multiple choice questions (MCQs) quiz in measuring the level of delirium care knowledge was found. The MCQs is a strictly objective instrument that has been widely used in examinations and knowledge tests internationally (Gabriel & Violato, 2009; Tweed, Purdie, & Wilkinson, 2020). Little is known about whether a useful examination instrument such as MCQs quiz is reliable to assess knowledge of delirium care. Therefore, this study reported the development and validation process of a MCQs quiz in examining the knowledge of delirium care among critical care nurses.

### 1.1 Background

As noted, clinical guidelines have provided the summary evidence of delirium care in terms of understanding the predisposing and precipitating risk factors, delirium risk prediction model, validated assessment tool, the short- and long-term outcomes of delirium in critically ill adults and so on (Devlin et al., 2018). Aforementioned available delirium knowledge instruments have covered these topics such as diagnosis, risk factors, sign and symptoms, assessment as well as the health outcomes of delirium (Elliott, 2014; Öztürk Birge et al., 2020). A growing body of research has highlighted the utilisation of a delirium prediction model in detecting delirium risk in ICU (Cowan, Preller, & Goudie, 2020; van den Boogaard et al., 2012; van den Boogaard et al., 2014). A recently meta-analysis also suggested that the delirium risk prediction model could be considered to applied in ICU settings (Ho, Chen, et al., 2020). Regarding the validated delirium assessment tools, a systematic review and diagnostic meta-analysis introduced several tools commonly used in ICU and indicated both the Confusion Assessment Method for the ICU (CAM-ICU) as well as the Intensive Care Delirium Screening Checklist (ICDSC) showed excellent sensitivity and specificity. Nevertheless, the CAM-ICU is recommended as the optimal tool due to its higher diagnostic test accuracy (Ho, Montgomery, et al., 2020). Considering to empower critical care nurses with sufficient level of delirium care knowledge and to be in line with the clinical guidelines (Department of Health and Ageing., 2011; Devlin et al., 2018; Scottish Intercollegiate Guidelines Network [SIGN], 2019), it is necessary to translate and implement latest evidence-based knowledge into clinical practice.

Given the rationale of employing a MCQs quiz in our study, several strategies to increase the quality of the MCQ can be considered in developing the quiz. For instance, 1) using the single best answer (SBA) which is the most common types of MCQs, so that the respondents would be familiar with the type and process of a test; 2) including allied distractors with correct answer which could increase the difficulty and discrimination of the questions; 3) adopting the use of “all/none of the above,” options can potentially increase the item difficulty because it is harder to ascertain other options are correct or incorrect; 4) designing a brief introduction stem and a lead-in question, respondents will be able to read and think the given stem to choose the most appropriate or likely option (Coughlin & Featherstone, 2017; Schuwirth & Van Der Vleuten, 2004). In order to develop a high quality MCQs quiz within the field of medical education, it is also essential to create questions with workplace simulated (i.e. a stem can be a clinical scenario description), problem solving and decision making descriptions, which can maximise the impacts and validity of the MCQ (Coughlin & Featherstone, 2017; Maguire, Skakun, & Triska, 1997; Naeem, van der Vleuten, & Alfaris, 2012). Currently, there is no validated MCQs quiz for testing the delirium care knowledge among critical care nurses. Therefore, this study aimed to design a MCQs quiz to examine delirium care knowledge based on the existing clinical guidelines and published literature and evaluate its psychometric properties. The specific research question is ‘whether the MCQs quiz is a reliable tool to assess delirium care knowledge among critical care nurses?’

## 2. METHODS

### 2.1 Aims

The purpose of the study was to develop and psychometrically test a MCQs-based delirium care knowledge quiz for critical care nurses.

### 2.2 Design

This reliability and validity study described a two-phase process in development and validation of the delirium care knowledge quiz.

### 2.3 Phase I: Development of the delirium care knowledge quiz

The delirium care knowledge quiz was designed by the research team based on 1) the ‘Delirium Care Pathways’ which pre-dated the national standards for delirium care in Australia (Department of Health and Ageing., 2011); 2) the PADIS 2018 guideline of delirium care in ICU setting in USA (Devlin et al., 2018); 3) SIGH evidence-based guideline, a national clinical guideline in Scotland (SIGN, 2019); and 4) the existing evidence including systematic review and meta-analyses of delirium assessment and detection (Ho, Chen, et al., 2020; Ho, Montgomery, et al., 2020). To assess the level of delirium care knowledge in critical care nurses, a 20-item pool was generated by the research team (Table 1). The content of the preliminary version of the delirium care knowledge quiz including signs and symptoms, risk factors and aetiologies, assessment and detection of delirium care. The single-select, multiple choice question (MCQ) was considered as the most suitable form of the delirium care knowledge quiz. Furthermore, MCQs was used in a number of national and healthcare-related specialty board examinations (Coughlin & Featherstone, 2017). Thus, our target respondents might not feel difficult to answer. In the initial quiz, most items (n=14) were 4-option MCQs, some were 2-option (n=4), and two questions were 5-option MCQs. A correct SBA was allocated 1 point while incorrect answer or missing response was scored 0 points, yielding the range of the quiz from 0 to 20 points from 20 initial items.

**Table 1.**
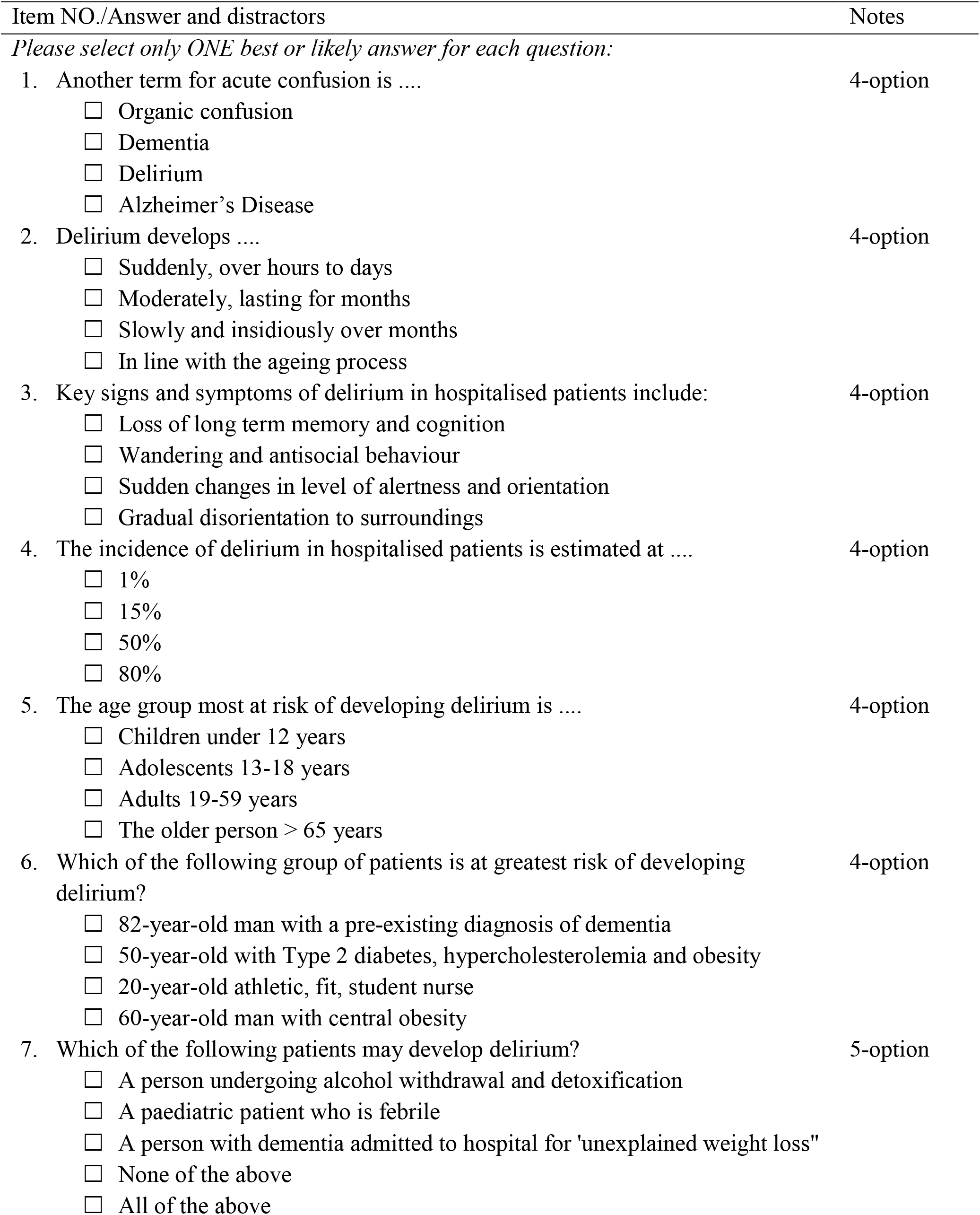
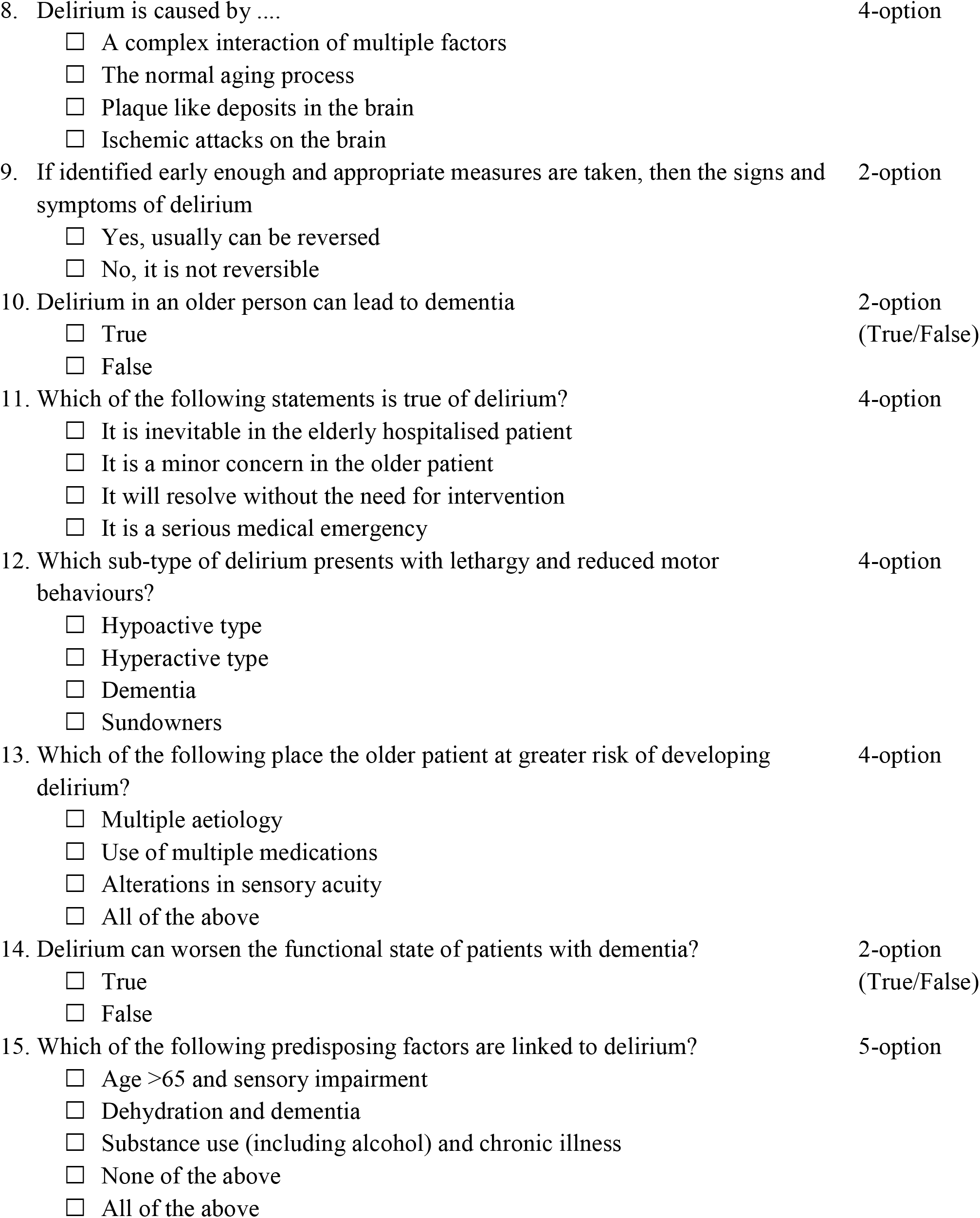
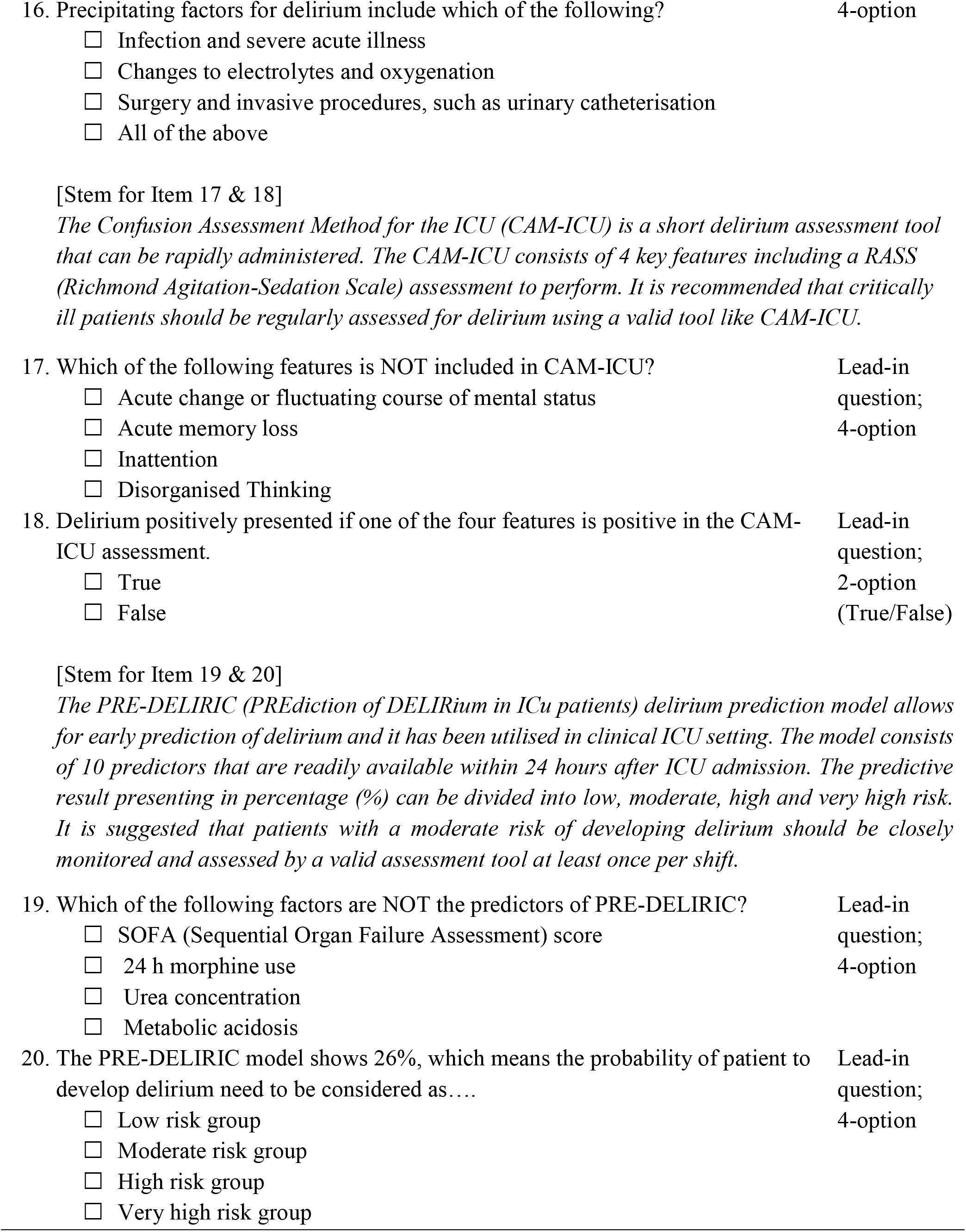
20-item pool of the initial delirium care knowledge quiz

#### 2.3.1 Content validity

The content of the quiz was examined by five experts consisted of an intensivist, two senior critical care nurses, a clinical nurse educator and a psychiatrist. The content agreement among experts in four aspects including the content relevance, applicability, representativeness, and clarity were evaluated using a 5-point Likert scale (1 = very low to 5 = very high) on each item (Haynes, Richard, & Kubany, 1995; Polit & Beck, 2006). The values of Content Validity Index (CVI) for the content relevance, applicability, representativeness, and clarity were 80.6%, 80.4%, 83.0%, and 87.6%, respectively. The CVI of each item was ≥ 80%, thus no question was eliminated and only one question was slightly amended in the wording for its presentation based on the comments from experts. The overall CVI calculated for the delirium care knowledge quiz was 82.9%.

#### 2.3.2 Face validity

Face validity refers to the appearance of an instrument (Considine, Botti, & Thomas, 2005). To assess the face validity, clarity and readability of the delirium care knowledge quiz, the 20-item pilot quiz was administered to 15 volunteer nurses with at least one-year clinical experience in ICU. Considering the target population is critical care nurses, the volunteer nurses were purposive invited by primary investigator from ICUs at three university-affiliated hospitals. All nurses reported that the quiz was understandable, and no further change was required.

#### 2.3.3 Sample for pilot testing and psychometric properties

Participants were recruited from three acute metropolitan teaching hospitals in northern Taiwan. Selection criteria were registered nurses who worked in ICU, and were older than 20 years. Critical care nurses who worked in the neonatal ICU or emergency room were excluded. The pilot quiz was distributed using a web-based survey tool (Qualtrics, Provo, UT) to gather information from the respondents. The online survey consisted of characteristics of respondents and the pilot quiz. Only basic characteristics information was collected including age, gender, ICU types (medical/surgical/mixed), and education level. The invitation with an electronic link with a QR code was sent to the department of nursing and the head nurses of ICUs. The title page of the online survey provided information regarding the study aim, the use and storage of data. Participants were also fully informed that this was an anonymous survey and all collected data were de-identified. Completion of the survey was considered to imply consent. In order to conduct the factor analysis, the estimated sample size for a 20-item test was 200 participants, which was based on the rule of 10 people per item (Goldberg & Velicer, 2006). The quiz was piloted with 228 critical care nurses. Eleven observations were dropped by primary investigator due to the missing responses were more than 60% of the whole quiz. Finally, data from 217 critical care nurses were analysed. Data was collected during 01 October 2020 to 07 November 2020.

#### 2.3.4 Item analysis

The item difficulty index (*P*) was generated based on the correct response rate for each item (Ahmann & Glock, 1981; Considine et al., 2005). It was used to describe the distribution of difficulty of a quiz. The ratio ranging from 0.00 to +1.00 was calculated by the number of the respondents with correct answer (*K*) divided the total number of the respondents. If the ratio of an item approaches +1.00 that means the question is easy. It is recommended that items with an item difficulty index <0.20 and >0.80 which implied the question is extremely easy or difficult should be considered to remove (Ahmann & Glock, 1981; Rush, Rankin, & White, 2016).

The item discrimination index was used to evaluate the discriminating degree of a quiz between respondents answering an item correctly or incorrectly (Considine et al., 2005; Rush et al., 2016). The following steps were conducted to compute the item discrimination index for each item: 1) sorted the total scores of the quiz in an ascending order in the statistics software, and ranked the first respondent with lowest score as from one; 2) defined high-scoring group and low-scoring group. The common method was 27%-70% rule proposed by Kelly (1939), the first 27% of respondents with lower scores and 70% of respondents with higher scores (Kelley, 1939) were identified (n=125). Thus, the 59 respondents (27% of 217, ranked 1-59) were marked as the low-scoring group and 66 (70% of 217, ranked 152-217) respondents were labelled as the high-scoring group; 3) calculated the correct response rate on each item in both high-scoring group (*PH*) and low-scoring group (*PL*); 4) performed an independent *t* test to examine the difference in total scores between high-scoring group and low-scoring group in order to confirm the significant differences between groups exist; Finally, the value of the discrimination index (*D*) for each item was computed according to the following formula:

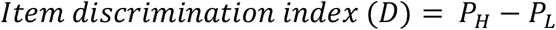

The index ranges from −1.00 to +1.00, and items with the index <0.25 should be deleted (Ebel & Frisbie, 1991). In this study, we removed items which suggested by item difficulty index and item discrimination index and finalised the delirium care knowledge quiz. The reliability test and psychometric evaluation were undertaken to examine the final quiz in the validation process.

### 2.4 Phase II: Validation of the delirium care knowledge quiz

#### 2.4.1 Internal consistency and test-retest reliability

The quiz of delirium care knowledge was finalised after the reduction of question items by item analysis. The internal consistency was calculated using the Kuder–Richardson Formula 20 (KR-20) coefficient to ensure the consistency of each item. Item-total correlations and correlated item-deleted analyses were also conducted to examine the robustness of the item reliability. Moreover, the final quiz was divided to two parts by odds- and even ordered and the split-half coefficient was computed to demonstrate the overall consistency. The Cronbach’s alpha or KR-20 coefficient which greater than 0.70 is considered as a reliable item/tool (Tavakol & Dennick, 2011).

The test-retest reliability reflects the variation in measurements taken by a tool on the same participants under the same conditions (Koo & Li, 2016). The intraclass correlation coefficient (ICC) was used to report the test-retest reliability of the quiz. Fifteen critical care nurses who took the 20–item pool test and assessed the face validly were invited again to undertake the finalised quiz (16 items) after 30 days. The data was compared and adopted to calculated the ICC. The values of ICC between 0.5 and 0.75 indicate moderate, 0.75-0.9 indicate good reliability, and values greater than 0.90 indicate excellent reliability (Portney & Watkins, 2009).

#### 2.4.2 Construct validity

For the contrast validity of the final quiz, firstly, the parallel analysis with a graphical scree plot approach was employed to determine the number of factors in this quiz. Then, the exploratory factor analysis (EFA) was applied to confirm the construct validity of the quiz. The Kaiser-Meyer-Olkin (KMO) measure of sampling adequacy (using a cut-off of 0.5), and Barlett’s Test of Sphericity (*p* < .001) were employed to ensure the appropriateness of data set for EFA (Kaiser, 1974). The eigenvalues and factor loadings were evaluated for the construct validity of the quiz. In the EFA, item with factor loading < 0.4 should be removed and re-modelling the structure of EFA (Hair, Black, Babin, & Anderson, 2010). The average communalities value between 0.5 and 0.6 is considered as acceptable for sample size around 200 (MacCallum, Widaman, Preacher, & Hong, 2001).

### 2.5 Ethical and considerations

Participants were voluntary and all data were anonymous and de-identified. The protocol of this cross-sectional pilot study was granted by a university Ethical Committee (Approval Number: N202009052).

### 2.6 Data analysis

The Statistical Package for Social Sciences (IBM SPSS Statistics for Windows, Version 25.0. Armonk, NY: IBM Corp.) was used to analysed the abovementioned data, reliability and validity tests. Frequency, percentage, mean, and standard deviation were used to present and distribution of variables in characteristics of nurses. All tests were two-tailed and the significance level (*α*) was set at 0.05.

## 3. RESULTS

The characteristics of the participants are shown in Table 2. Most of the critical care nurses were female (87.5%), worked in medical (45.2%) or surgical (44.7%) ICU, with an undergraduate degree (93.1%). The mean age of nurses was 31.2 years (SD = 7.1), 57.9% (n=124) were aged 20 to 30 years, 30.8% (n=66) were 31-40 years, and 11.2 % (n=24) were over 40 years.

**Table 2.** Characteristics of participants in pilot testing (*N* = 217)

| Characteristics | Mean (SD), range | n (%) |
| --- | --- | --- |
| Age, years (n=190) | 31.2 (7.1), 21-58 |  |
| 20-30 |  | 124 (57.9) |
| 31-40 |  | 66 (30.8) |
| 40+ |  | 24 (11.2) |
| Gender |  |  |
| Female (n=216) |  | 189 (87.5) |
| Male |  | 27 (12.5) |
| Type of ICU |  |  |
| Medical |  | 98 (45.2) |
| Surgical |  | 97 (44.7) |
| Mixed |  | 22 (10.1) |
| Education level |  |  |
| Undergraduate |  | 202 (93.1) |
| Postgraduate |  | 15 (6.9) |

*Note.* SD = standard deviation; ICU = intensive care unit.

### 3.1 Phase I: Finalise the quiz

#### 3.1.1 Item analysis

The independent *t* test showed there was a significant difference in total scores between high-scoring (n=66) group and low-scoring (n=59) group (*t*=-25.908, *p*<.001) that supported an excellent discrimination of the quiz. The item difficulty index (*P*) indicated item 1 and 10 were too easy and item 4 and 11 were too difficult in the quiz (out of the 0.20-0.80 range), and all item discrimination indices (*D*) for these four items were <0.25, which also demonstrated low discrimination of the questions (Table 3). Accordingly, item 1, 4, 10, 11 were removed from the quiz. Table 3 also presented the difficulty indices of the quiz ranging from 34.6 to 79.7 within the remained items. The discrimination indices in two items were greater than 0.40 which showed these questions were very good discriminator in the quiz. The final quiz with 16-item was confirmed and finalised. The mean score of the delirium care knowledge quiz on the 16-item MCQs was 10.3 (SD = 4.0) with a range 1-16.

**Table 3.** Results of item analysis on pilot quiz (*N* = 217)

| 20-item pool | $K$ | $P$ | $P_H$ (n=66) | $P_L$ (n=59) | $D$ | Item remaining |
| --- | --- | --- | --- | --- | --- | --- |
| 1 | 193 | 88.9 | 0.512 | 0.344 | 0.168 | Removed |
| 2 | 170 | 78.3 | 0.488 | 0.176 | 0.312 | ✓ |
| 3 | 123 | 56.7 | 0.512 | 0.104 | 0.408 | ✓ |
| 4 | 42 | 19.4 | 0.168 | 0.040 | 0.128 | Removed |
| 5 | 167 | 77.0 | 0.520 | 0.176 | 0.344 | ✓ |
| 6 | 166 | 76.5 | 0.496 | 0.176 | 0.320 | ✓ |
| 7 | 143 | 65.9 | 0.528 | 0.112 | 0.416 | ✓ |
| 8 | 173 | 79.7 | 0.520 | 0.216 | 0.304 | ✓ |
| 9 | 168 | 77.4 | 0.520 | 0.224 | 0.296 | ✓ |
| 10 | 204 | 94.0 | 0.512 | 0.416 | 0.096 | Removed |
| 11 | 40 | 18.4 | 0.144 | 0.032 | 0.112 | Removed |
| 12 | 154 | 71.0 | 0.528 | 0.152 | 0.376 | ✓ |
| 13 | 162 | 74.7 | 0.512 | 0.136 | 0.376 | ✓ |
| 14 | 62 | 28.6 | 0.312 | 0.008 | 0.304 | ✓ |
| 15 | 172 | 79.3 | 0.528 | 0.256 | 0.272 | ✓ |
| 16 | 171 | 78.8 | 0.528 | 0.248 | 0.280 | ✓ |
| 17 | 97 | 44.7 | 0.360 | 0.048 | 0.312 | ✓ |
| 18 | 75 | 34.6 | 0.368 | 0.016 | 0.352 | ✓ |
| 19 | 91 | 41.9 | 0.360 | 0.080 | 0.280 | ✓ |
| 20 | 134 | 61.8 | 0.488 | 0.136 | 0.352 | ✓ |

Note. $K$ = number of correct response; $P$ = item difficulty index; $P_H$ = correct response rate in high-scoring group; $P_L$ = correct response rate in low-scoring group; $D$ = item discrimination index.

### 3.2 Phase II: Evaluate the reliability and validity of the final quiz

#### 3.2.1 Internal consistency and test-retest reliability

The total quiz had an internal consistency of 0.854 (KR-20) which indicated the final quiz is a reliable test. The Spearman-Brown coefficient of 0.767 for split-half reliability also confirmed the reliability of the final quiz. The item-total correlations revealed that all items were correlated with the total score of the quiz significantly (*p*<0.0001). The results of correlated item-total correlation and the Cronbach’s alpha if item deleted were summarised in Supplementary Table 2. The ICC of 0.961 (95% CI: .903-.989, *p*<0.0001) highlighted an excellent test-retest reliability of the quiz.

#### 3.1.2 Construct validity

In Figure 1, four factor components were determined by the parallel analysis as the eigenvalue of the fifth factor was less than 1.0 in the graphical scree plot. The EFA using principal component analysis with varimax rotation was performed. The KMO of 0.823 (>0.5) and the Barlett’s Test of Sphericity (*p* < 0.0001) suggested the collected data was appropriate to conduct EFA. In the EFA, the eigenvalue of four factors could explain the total variance of 60.87%. All factor loadings of items were >0.439 or above. The average communality value was 0.653, and in each item was >0.4, indicating each item shares some common variance for all 16 items (Table 4).

**Table 4.**
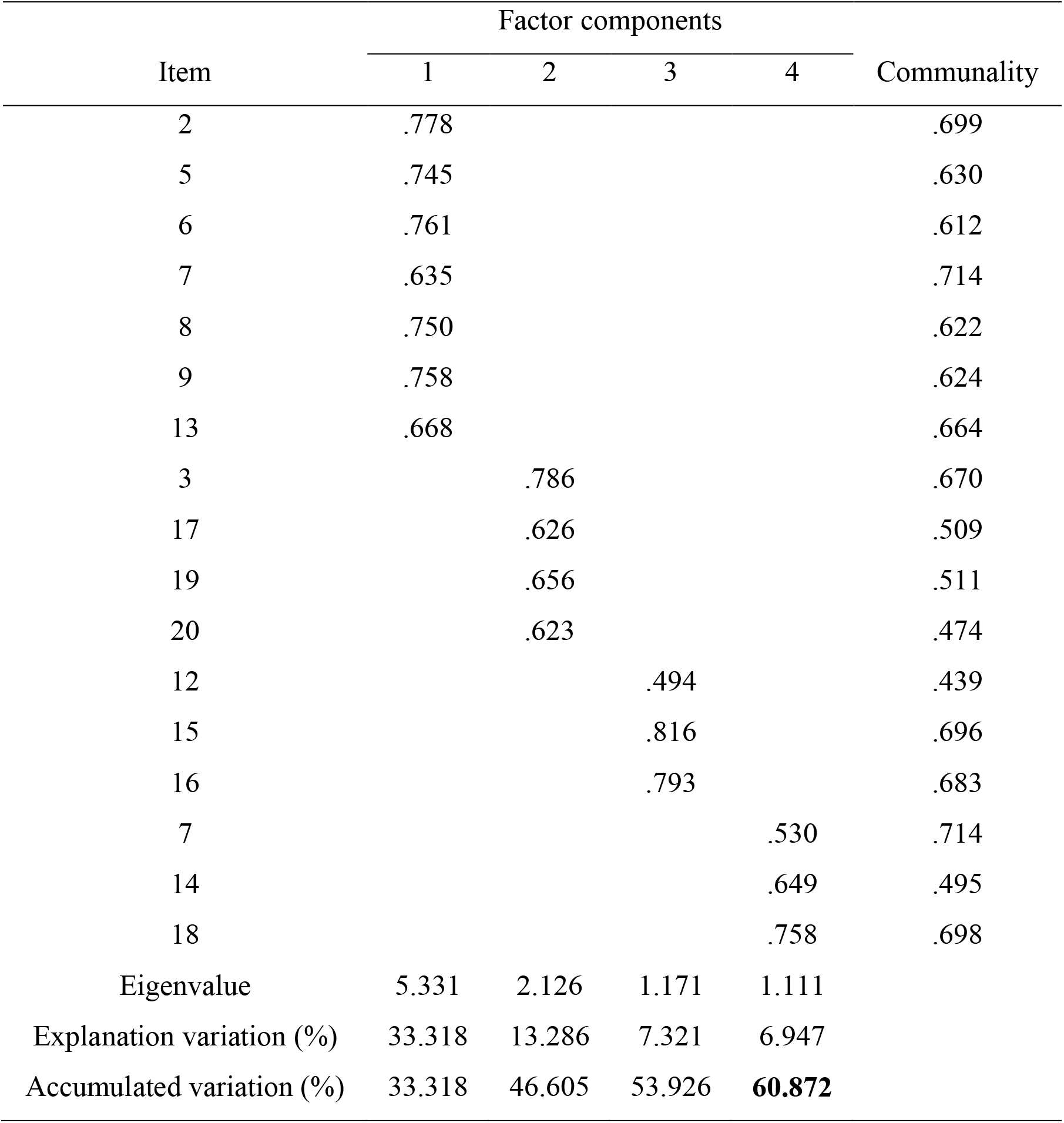
Factor loadings and eigenvalues from the exploratory factor analysis (*N* = 217)

| Item | Factor components |  |  |  | Communality |
| --- | --- | --- | --- | --- | --- |
|  | 1 | 2 | 3 | 4 |  |
| 2 | .778 |  |  |  | .699 |
| 5 | .745 |  |  |  | .630 |
| 6 | .761 |  |  |  | .612 |
| 7 | .635 |  |  |  | .714 |
| 8 | .750 |  |  |  | .622 |
| 9 | .758 |  |  |  | .624 |
| 13 | .668 |  |  |  | .664 |
| 3 |  | .786 |  |  | .670 |
| 17 |  | .626 |  |  | .509 |
| 19 |  | .656 |  |  | .511 |
| 20 |  | .623 |  |  | .474 |
| 12 |  |  | .494 |  | .439 |
| 15 |  |  | .816 |  | .696 |
| 16 |  |  | .793 |  | .683 |
| 7 |  |  |  | .530 | .714 |
| 14 |  |  |  | .649 | .495 |
| 18 |  |  |  | .758 | .698 |
| Eigenvalue | 5.331 | 2.126 | 1.171 | 1.111 |  |
| Explanation variation (%) | 33.318 | 13.286 | 7.321 | 6.947 |  |
| Accumulated variation (%) | 33.318 | 46.605 | 53.926 | **60.872** |  |

**Figure 1.**
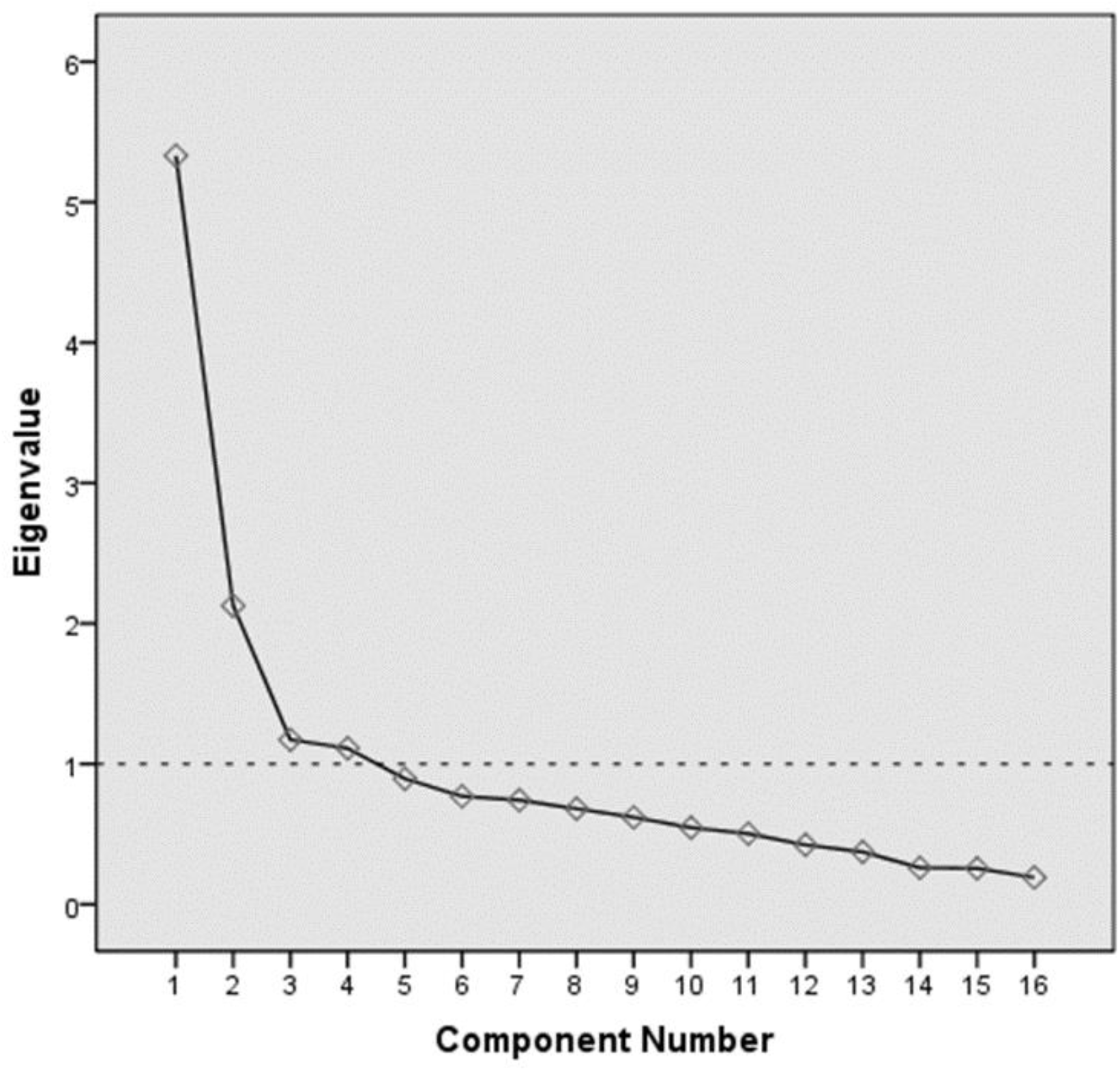
Parallel analysis (scree plot) for determining the number of the factors

## 4. DISCUSSION

This study aimed at developing and psychometrically testing the 16-item MCQs quiz of the delirium care knowledge. The results have demonstrated our MCQs quiz is a reliable and validated tool to assess delirium care knowledge among critical care nurses. Among all items in the quiz, one difficult item with good discrimination degree is about interpreting the result of CAM-ICU assessment (item 18). Surprisingly, this item had only 34.6% correct response rate from critical care nurses, which highlighted an urgent need to educate and improve their knowledge of using CAM-ICU. A possible explanation is that participants are more familiar with other delirium assessment tools such as ICDSC or 4AT in their daily practice. Still, critical care nurses should be able to recognise the correct answer because the CAM-ICU had been introduced and suggested to be used in the Taiwan Clinical Practice Guidelines for the Management of Pain, Agitation, and Delirium in Adult Intensive Care Unit (Taiwan PAD) since 2016 (Tay, Chan, Chen, Chou, & Huang, 2016). Consistent with findings from previous study conducted in the UK, education on ICU delirium is warranted in particular the training in delirium screening (Elliott, 2014). Thus, there is a growing demand for implementing education interventions and programs to improve critical care nurses’ knowledge in CAM-ICU assessment.

The fundamental content of the items including signs and symptoms, risk factors of delirium are important and similar items could be also found in previous tools in assessing delirium knowledge (Elliott, 2014; Öztürk Birge et al., 2020). However, our findings indicated that these items testing basic knowledge (item 8, 13, 15, 16) seem to be too easy (item difficulty index >70%). These results are line with previous study using a validated tool (Öztürk Birge et al., 2020). Although these items were not removed, future studies may consider revising the options and adding distractors to ensure the concept of the knowledge is measured, and reduce the chance of correct response rate by guessing. For example, using either ‘None of the above’ or ‘All of the above’ rather than using both in a single question which might prompt the correct answer to respondents (Rush et al., 2016).

Our findings also discovered that items with stem and lead-in questions work well in terms of the response rate and item difficulty. None of missing responses were received from these four items and the difficulty were considered acceptable (item difficulty index between 34.6-61.8%). In our quiz, we did not design a case report-like stem in our item (i.e. An 83-year-old woman presented with confusion…) due to the consideration in length of the quiz while it is recommended to be included (Considine et al., 2005; Coughlin & Featherstone, 2017; Naeem et al., 2012). This is an interesting topic for future studies to discover how the MCQs with clinical scenario description can extend the knowledge measurement beyond testing simple concept of knowledge.

Understanding the sufficient level of knowledge depends on a reliable and valid tool. The findings indicated that this MCQs quiz has appropriate psychometric properties. Although this quiz was developed for and validated in critical care nurses, we recommended that this quiz could be also used in undergraduate curricula as an examination quiz and assessment task in critical care nursing related subjects. The reason for adopting this quiz is because components were developed according to clinical guidelines which provided the practical guidance that should be integrated into the learning objectives. Central to the learning outcomes in delirium care among critical care nurses or nursing students is the clinical practice driven by the knowledge acquisition.

### 4.1 Limitations

This study had several strengths such as developing the first validated MCQs quiz that can be used in both undergraduate curricula and continuing medical/nursing education, reporting the estimates of reliability and validity comprehensively, and adopting strategies to create high quality MCQs and using item analysis to test the question difficulty and discrimination. Nevertheless, some limitations were noted in this study. First, in this self-developed instrument, we did not have theoretical perspectives to support the development of the quiz. In order to design a knowledge-based assessment instrument, we used more than one clinical practice guidelines and the latest evidence-based literature to create initial items. The concept of delirium care knowledge was included in our quiz, and the multidimensional psychometric properties were carefully estimated in the validation process. Second, the generalisability/external validity of the results might be limited due to the all participants were recruited from a single area, northern Taiwan. Lastly, the self-report online survey method might cause the social-desirability bias, for example, respondents can search the correct answer before response to the online survey. Despite these limitations, the quiz had the optimal statistical properties with regard to measuring the level of delirium care knowledge among critical care nurses.

## 5. CONCLUSION

Understanding the level of delirium care knowledge among critical care nurses is vital to support future education interventions in ICU settings. The MCQs quiz developed in this study is a reliable and valid tool that can be implemented to assess the level of delirium care knowledge. It is an evidence-based quiz designed for future research and education purposes in delirium care that has significant implications for knowledge test by using MCQs in clinical practice.

## Supporting information

Reporting guideline

## Data Availability

Non-digital data available

