## Supplementary Table 1 for "Development and validation of a multiple-choice question-based delirium care knowledge quiz for critical care nurses"

**Supplementary Table 1.** Item-Total correlation

| Item | Correlated item-Total correlation | Cronbach's alpha if item deleted |
| --- | --- | --- |
| 2 | .608 | .839 |
| 3 | .494 | .845 |
| 5 | .620 | .838 |
| 6 | .578 | .841 |
| 7 | .530 | .843 |
| 8 | .570 | .841 |
| 9 | .532 | .843 |
| 12 | .507 | .844 |
| 13 | .673 | .836 |
| 14 | .341 | .852 |
| 15 | .325 | .852 |
| 16 | .403 | .849 |
| 17 | .323 | .854 |
| 18 | .411 | .849 |
| 19 | .329 | .854 |
| 20 | .458 | .846 |
